# Inactivation of DRG1, encoding a translation factor GTPase, causes a Recessive Neurodevelopmental Disorder

**DOI:** 10.1101/2022.09.20.22279914

**Authors:** Christian A. E. Westrip, Franziska Paul, Fathiya Al-Murshedi, Hashim Qaitoon, Breana Cham, Sally C. Fletcher, Eline Hendrix, Uncaar Boora, Alvin Yu Jin Ng, Carine Bonnard, Maryam Najafi, Salem Alawbathani, Imelda Lambert, Gabriel Fox, Byrappa Venkatesh, Aida Bertoli-Avella, Ee Shien Tan, Almundher Al-Maawali, Bruno Reversade, Mathew L. Coleman

## Abstract

DRG1 is a highly conserved member of a class of GTPases implicated in ribosome biogenesis and translation. The expression of mammalian DRG1 is elevated in the central nervous system during development, and its function has been implicated in fundamental cellular processes including protein synthesis and cellular proliferation. Using exome sequencing, we identified rare and likely pathogenic germline *DRG1* variants including three stop-gained p.Gly54*, p.Arg140*, p.Lys263* and a p.Asn248Phe missense variant. These alleles segregate recessively in four affected individuals from three unrelated families and cause a neurodevelopmental disorder with global developmental delay, microcephaly, short stature and craniofacial anomalies. Using functional assays, we show that these loss-of-function variants: 1) severely disrupt DRG1 mRNA/protein stability in patient-derived fibroblasts, 2) impair it’s GTPase activity *in vitro* and 3) compromise it’s binding to partner protein ZC3H15. Consistent with the importance of DRG1 in humans, targeted inactivation of *Drg1* in mice resulted in pre-weaning lethality. Our work highlights the importance of DRG1 GTPase activity for normal development and underscores the significance of translation factor GTPases in human physiology and homeostasis.

## INTRODUCTION

GTPases are a large enzyme superfamily with critical roles in fundamental cellular processes (Sahai & Marshall, 2002; Wennerberg *et al*, 2005; Verstraeten *et al*, 2011). Central to their function is the ability to bind and hydrolyse GTP (Bourne *et al*, 1991; Vetter & Wittinghofer, 2001; Wittinghofer & Vetter, 2011), which confers the ability to cycle between activity states and to act as molecular ‘switches’.

The GTPase family consists of two subgroups referred to as SIMIBI and TRAFAC (Leipe *et al*, 2002). The TRAFAC group was named after members that function as Translation Factors, but also includes RAS and heterotrimeric GTPases (Leipe *et al*, 2002). Less well-characterised TRAFAC GTPases include OBG-(spoOB-associated GTP-binding protein) and HflX-(high frequency of lysogenization protein X) like GTPases. OBG/HflX GTPases are an ancient class of enzymes with some members present in all domains of life (Leipe *et al*, 2002). OBG/HflX GTPases have roles in ribosome regulation/biogenesis, translation or RNA binding (Daugeron *et al*, 2011; Kallstrom *et al*, 2003; Zhang & Haldenwang, 2004).

The developmentally-regulated GTP-binding (DRG) proteins, DRG1 (MIM603952) and DRG2 (MIM602986), are highly conserved OBG/HflX GTPases (Li & Trueb, 2000; Westrip *et al*, 2021) that interact with RNA and ribosomes, consistent with a proposed translational role (Ishikawa *et al*, 2003, 2009; Daugeron *et al*, 2011). Indeed, structural analysis of DRG1 places it within the large ribosomal subunit where it is required for relieving ribosomal pausing (Zeng *et al*, 2021). DRGs have also been studied in other contexts (Westrip *et al*, 2021). DRG1 expression is elevated during development of the central nervous system (Kumar *et al*, 1992; Sazuka *et al*, 1992; Ishikawa *et al*, 2005; Wei *et al*, 2004). It is also required for cell proliferation and has been implicated in cancer-associated processes and tumourigenesis (Lu *et al*, 2016; Ling *et al*, 2020; Kiniwa *et al*, 2015; Jiang *et al*, 2016). The importance of these functions and their potential roles in physiology and disease remain unclear however.

Whilst mutation of genes encoding small GTPases such as Ras have been widely studied in the context of cancer (Sahai & Marshall, 2002) and, more recently, neurodevelopmental disorders (Shieh, 2019), the role of the wider TRAFAC GTPase family is much less well understood. To date there have been no pathogenic germline variants identified in genes of the OBG/HflX GTPase family, including the DRG GTPases. Since gene expression control at the level of translation is now recognised as an increasingly important area of deregulation in inherited disease (Scheper *et al*, 2007), further studies of these enigmatic GTPases is warranted.

Here, we identify inherited loss-of-function variants in *DRG1* in three pedigrees that present with a novel developmental disorder associated with global developmental delay, failure to thrive, microcephaly, and craniofacial dysmorphism. We show that the disease variants severely damage DRG1 protein expression, interactions, and GTPase activity.

## RESULTS AND DISCUSSION

### Three families segregating recessive DRG1 variants

We report four individuals from three independently identified families with biallelic deleterious *DRG1* variants resulting in a neurodevelopmental syndrome (Figure 1A-B). With the exception of Family 2 (F2, 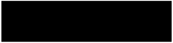 ancestry), the other families were of 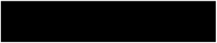 background and consanguineous (F1 and F3). The phenotypes in these 4 individuals consist of a global developmental delay, failure to thrive, microcephaly, intellectual deficit, and craniofacial anomalies. All four patients presented with intrauterine growth retardation (IUGR) at birth, and they continued to show significant growth delay. They showed a delay in attaining developmental milestones, but in general, they were able to walk and interact with their surroundings. They all had a variable speech delay. Detailed clinical descriptions and facial dysmorphism information is presented in Table 1 and Figure 1; a Clinical Report may be requested from the authors.

**Table 1.**
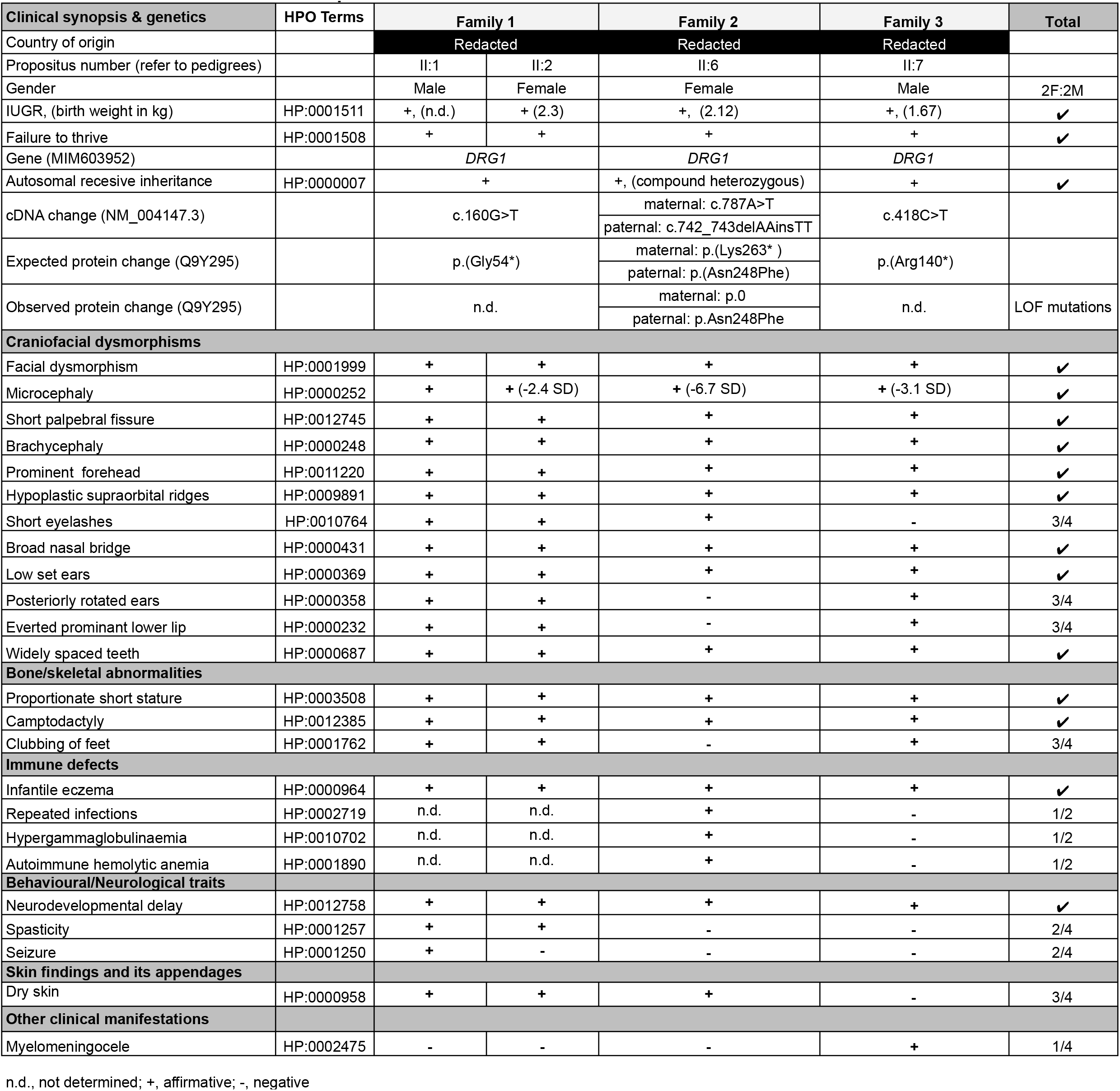
Clinical characteristics of four patients with biallelic DRG1 variants.

**Figure 1.**
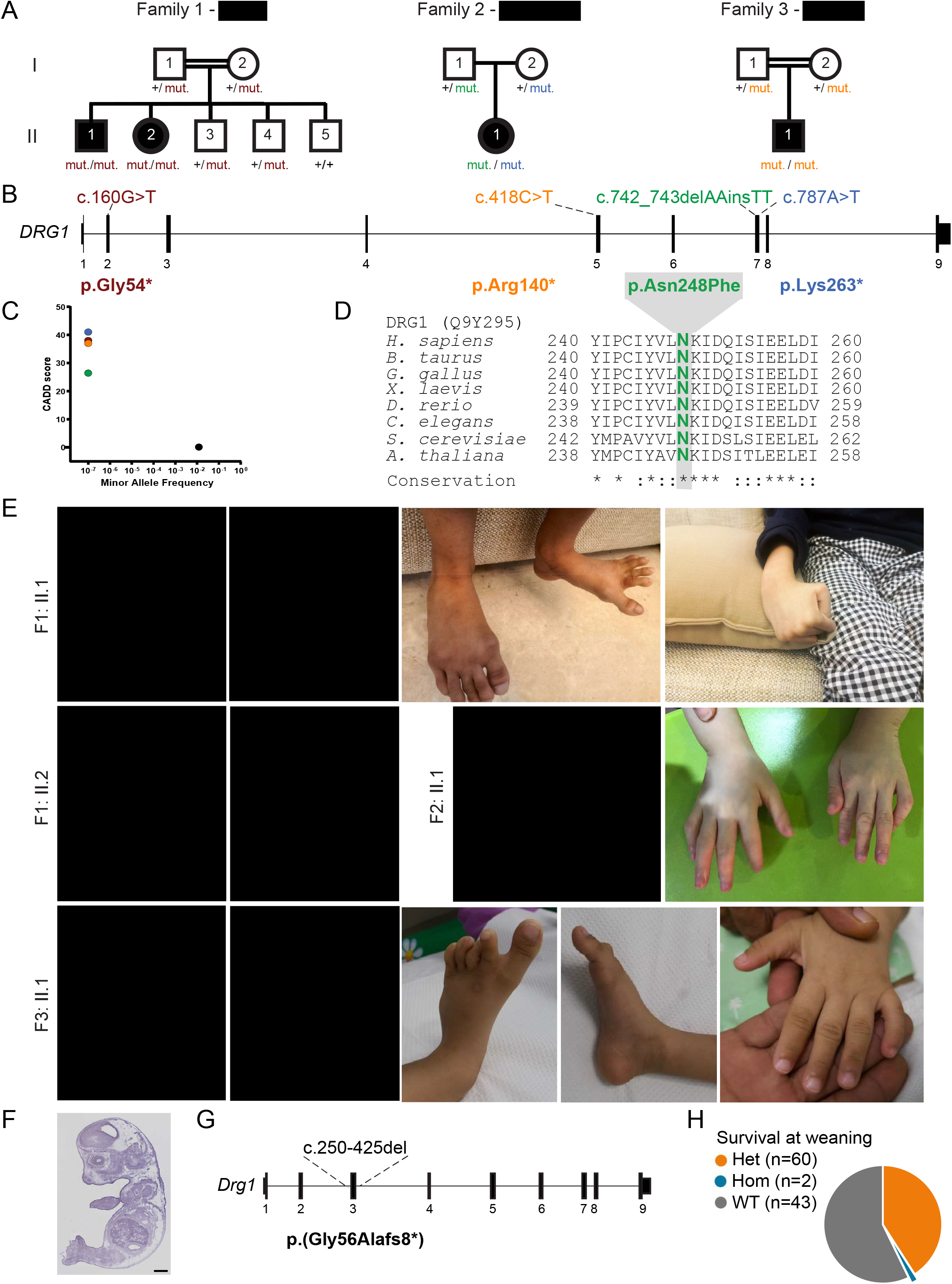
Three families segregating recessive *DRG1* loss-of-function variants. A. Pedigrees of three families in which affected children inherited recessive *DRG1* pLoF variants. Sanger segregation analysis of these germline variants is shown. B. Structure of the *DRG1* transcript indicating the location of the genomic variants (above) and their corresponding change in amino acid sequence (below). Variants are color-coded according to panel a. C. Minor allele frequency (MAF) and combined annotation-dependent depletion (CADD) score of homozygous DRG1 coding variants found in gnomAD v.2.1.1 (black dots) and those found in each family (color-coded dots). DRG1 is intolerant of genetic variation. D. p.Asn248Phe is located in a highly conserved region. Functionally conservative amino acid changes are indicated (* and:). E. Photographs of four affected children showing facial dysmorphism and camptodactyly, clubbed feed and eczema for selected patients. F. Ubiquitous *Drg1* expression in E14.5 mouse embryos by RNA *in situ* hybridization. Taken from the EMAGE gene expression database (Richardson *et al*, 2014) (http://www.emouseatlas.org/emage/; EMAGE:31607 June 2022). Scale bar 1 mm. G. Structure of the mouse *Drg1* transcript indicating the site of deletion in Drg1 KO mice. This deletion leads to a frameshift and premature stop codon within exon 4. H. Survival of *Drg1* KO mice upon weaning. This is significantly different from the expected litter distribution of 25% WT, 50% Het, 25% KO (Chi-square test χ^2^ (2, N = 105) = 34, p = 3.8×10^−08^).

According to genomAD (v2.1.1 and v3.1) no homozygous damaging variants have been reported for *DRG1*. None of the four germline variants identified through exome sequencing (p.Gly54*, p.Asn248Phe, p.Lys263* and p.Arg140*) have been reported in public databases (gnomAD, BRAVO/TOPmed, ExAC, 1000G) or in combined in-house databases consisting of >50,000 exomes/genomes. All three truncating variants were predicted to be deleterious, with combined annotation-dependent depletion (CADD) scores above 30 (Figure 1C). The missense p.Asn248Phe variant identified in case II:1 from Family 2 is located in a highly conserved region (Figure 1D, Figure S1) and thus annotated as a possible loss-of-function by MutationTaster with a computed CADD value of 26.4.

The *DRG1* gene has a residual variation intolerance score (RVIS) of −0.19 (placing it in the top 40% of human genes most intolerant to genetic variation) and a pLoF observed/expected score of 0.23 (gnomAD). This suggests that *DRG1* is a target of strong negative selection, which may be consistent with an essential function. Consistent with this, murine *Drg1* is ubiquitously expressed at E14.5 (Figure 1F) and essential for proper development: *Drg1* KO leads to a significantly lower survival rate at weaning age, with less than 2% *Drg1*^-/-^ pups obtained from heterozygous crosses (χ^2^ (2, N = 105) = 34, p = 3.8×10^−8^; Figure 1G-H).

Overall, these clinical and genetic findings suggest that homozygosity for pLOF variants is exceedingly rare in the general population and that the *DRG1* variants observed in these three kindreds are probably deleterious, most likely revealing the genetic etiology for this heretofore unknown syndrome.

Next, we experimentally investigated the impact of the *DRG1* variants described above. The p.Asn248Phe variant is located in the highly conserved GTPase domain (Figure 2A-B, Figure S1). Analysis of the primary sequence (Figure 1D) and tertiary structure (Figure 2B) indicates that Asn248 is completely conserved and is part of the G4 motif (<u>N</u>KID), which is required for binding to the guanine base of GTP (Bourne *et al*, 1991; Wittinghofer & Vetter, 2011). Mutation of the G4 motif is known to inhibit GTP binding and hydrolysis (Walter *et al*, 1986). Furthermore, substitution of Asparagine for the larger and more hydrophobic Phenylalanine at position 248 could also disrupt structural conformation beyond the GTPase domain (Figure 2B, explored in more detail below).

**Figure 2.**
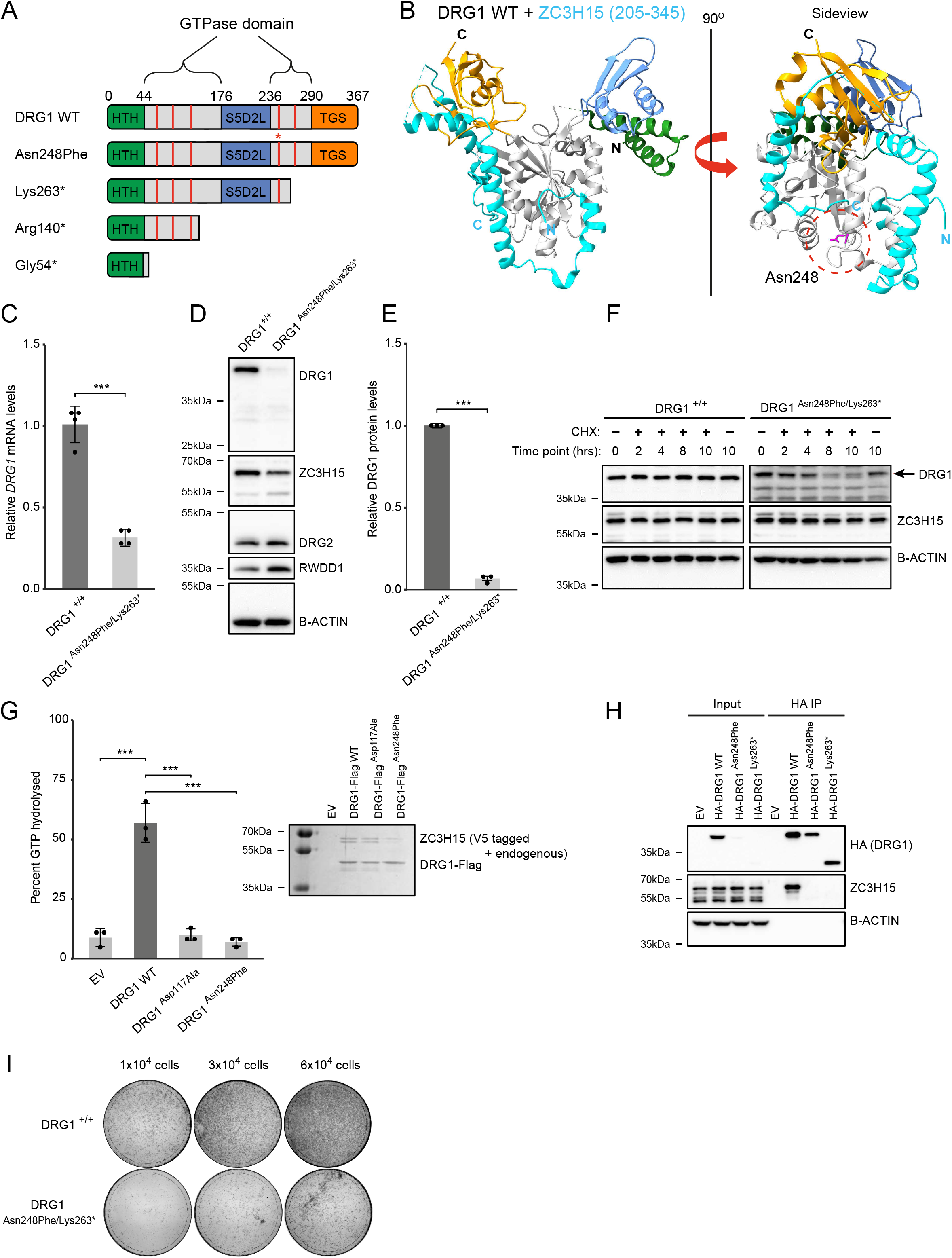
DRG1 variants reduce expression, GTPase activity and ZC3H15 binding. A. Domain organisation for wildtype (top) and mutant DRG1 proteins. Red bars indicate the five G motifs conserved in DRG GTPases. The location of the Asn248Phe variants is indicated with an asterisk. HTH: Helix Turn Helix, S5D2L: Ribosomal protein S5 domain 2-like domain, TGS: <u>T</u>hrRS, <u>G</u>TPase, and <u>S</u>poT domain. B. Structure of Rbg1 (yeast DRG1) showing the location of Asn248 and its proximity to the ZC3H15 binding interface. GTPase domain: grey, HTH: green, S5D2L: blue, TGS: orange. The C-terminal fragment of yeast ZC3H15 (Tma46) is also shown in cyan. C. Quantification of DRG1 mRNA levels in WT and DRG1 (Asn248Phe/Lys263*) mutated fibroblasts using RT-qPCR. Results are normalised to a GAPDH control. The data represents the mean with error bars showing the standard deviation of 4 biological repeats (data points shown). Statistical significance was estimated using a two-sample t-test. D. Western blots using protein extracts from wildtype (WT) and DRG1 Asn248Phe/Lys263* fibroblasts. E. Quantification of DRG1 protein levels relative to B-Actin. Data represents mean with standard deviation. Statistical significance was estimated using a two-sample t-test. F. Western blots of CHX stability assay with the DRG1 WT and DRG1 Asn248Phe/Lys263* fibroblasts. Cells were treated with 50μg/ml CHX and then harvested at the indicated time points. A 10hrs dimethyl sulfoxide (DMSO) control was also included. G. GTPase assay using C-terminally flag tagged DRG1 WT, Asp117Ala (positive control predicted to have no GTPase activity) and Asn248Phe that were co-transfected with C-terminally V5 tagged ZC3H15 in HEK293T cells and purified using anti-flag pulldown. Coomassie stained gel of purified DRG1/ZC3H15 complexes shown in inset. The data represents the mean with error bars showing the standard deviation of n=3 biological repeats (data points shown). Statistical significance was confirmed using a one-way ANOVA with Tukey HSD to estimate p values. H. N-terminal HA-tagged DRG1 wildtype (WT), Asn248Phe and Lys263* variants were transiently expressed in HeLa cells. Cell lysates were used in an anti-HA pulldown experiment followed by western blotting for the indicated proteins using input and pulldown samples. I. Colony forming assay using DRG1 WT and Asn248Phe/Lys263* fibroblasts. Cells were seeded on 10 cm plates then stained with crystal violet after 10 days. We note that these primary fibroblasts do not form compact colonies with clearly defined borders.

Three out of the variants are nonsense mutations, which likely trigger nonsense-mediated mRNA decay (NMD). Structural analysis indicates that translation of any residual mRNA would produce a severely truncated protein p.Gly54* that lacks essential functional domains (Francis *et al*, 2012) including TGS, S5D2L and GTPase domains (Figure 2A). The p.Arg140* variant would also result in a severely truncated protein lacking the TGS and S5D2L domains and half of the GTPase domain (Figure 2A and Figure S2A). A p.Lys263* DRG1 protein would lack part of the GTPase domain and the entire TGS domain (Figure 2A and Figure S2B). Importantly, in yeast, DRG1 requires the TGS domain to bind to ZC3H15 (also known as DFRP1, MIM619704) and for recruitment of DRG1/ZC3H15 complexes to polysomes (Francis *et al*, 2012). Importantly, ZC3H15 binding is also critical for DRG1 protein stability and GTPase activity (Ishikawa *et al*, 2005; Pérez-Arellano *et al*, 2013). Overall, the structural analyses strongly suggest that all four DRG1 variants are likely to impair GTPase activity and ZC3H15 binding, consistent with a likely loss of function.

To explore the functional consequences of the variants, we first expressed epitope-tagged DRG1 vectors in HeLa cells. Whereas HA-DRG1^Gly54*^ and HA-DRG1^Arg140*^ proteins were not expressed, the HA-DRG1^Lys263*^ and HA-DRG1^Asn248Phe^ proteins were detected, albeit with significantly reduced expression (Figure S2C). These data indicate that all four variants are likely deleterious to normal DRG1 expression. To validate this at the endogenous level, we cultured primary dermal fibroblasts from patient II-6 (p.Lys263*/p.Asn248Phe). Consistent with NMD of the endogenous DRG1^Lys263*^ transcript, we observed significantly reduced *DRG1* mRNA in the patient cells (Figure 2C). Sanger sequencing of *DRG1* cDNA only detected the p.Asn248Phe variant and no Lys263* (Figure S2D), suggesting that the residual transcript being the p.Asn248Phe variant. Fitting with the mRNA analysis, full-length endogenous DRG1 protein was dramatically reduced in patient cells (Figure 2D-E), and we were unable to detect a DRG1 protein species consistent with p.Lys263* (see Figure S2C for antibody validation). Overall, these data indicate that the p.Asn248Phe and p.Lys263* variants seriously impair normal DRG1 expression. Consistent with reciprocal regulation of DRG1 and ZC3H15 expression (Ishikawa *et al*, 2013), we observed a modest reduction in ZC3H15 protein in the patient cells (Figure 2D and Figure S2E). Importantly, these effects on DRG1/ZC3H15 were specific, as we did not observe reduced levels of DRG2 or its obligate binding partner RWDD1 (also known as DFRP2, Figure 2D, Figure S2F-G).

We next sought to better understand the impact of the p.Asn248Phe variant on DRG1 protein stability. Because we observed reduced protein expression from a heterologous promoter (Figure S2C) we postulated that this variant negatively regulates protein stability. Therefore, we performed cycloheximide-based protein turnover assays in transfected HeLa cells: The half-life of HA-tagged DRG1 protein was reduced from ∼5 hours in the wildtype to ∼1 hour for the HA-DRG1^Asn248Phe^ variant (Figure S2H). Importantly, we observed an even more dramatic effect on the stability of the endogenous protein in patient-derived cells (Figure 2F and Figure S2I). Overall, these results confirm that the p.Asn248Phe variant causes enhanced protein turnover and thus reduced DRG1 protein levels.

Since residual DRG1^Asn248Phe^ protein is expressed in the patient cells (Figure 2D), we next tested the impact on its enzymatic function. Therefore, we purified overexpressed HA-DRG1, HA-DRG1^Asn248Phe^, or HA-DRG1^Asp117Ala^ (a known inactivating mutation) from HEK293T cells, before analyzing GTPase activity (Figure 2G). Importantly, GTP hydrolysis catalyzed by the DRG^Asn248Phe^ variant was undetectable. For completeness, we also tested the p.Lys263* variant since it retains the bulk of the GTPase domain (Figure 2A). Interestingly, partially purified DRG1^Lys263*^ was also unable to support GTPase activity (Figure S3). Together, these data suggest that a second consequence of the p.Asn248Phe and p.Lys263* variants is loss of GTPase activity.

Interestingly, visual analyses of purified DRG1 (Figure 2G and Figure S3) suggested that these variants may also reduce ZC3H15 binding. To independently test this, we immunoprecipitated wildtype or variant HA-DRG1 from HeLa cells before western blotting for endogenous ZC3H15 (Figure 2H). Importantly, both p.Asn248Phe and p.Lys263* variants were unable to bind ZC3H15. Considering the importance of ZC3H15 binding for DRG1 activity and stability (Ishikawa *et al*, 2005, 2013; Francis *et al*, 2012), the effect of these variants on the complex may partly explain the loss of function observed. Overall, our combined functional analyses demonstrate that these novel patient variants severely impact the expression, GTPase activity and ZC3H15 binding of DRG1. Consistent with this, and the essentiality of DRG1, we find that patient-derived cells show a survival deficit in colony formation assays (Figure 2I).

In summary, we have found three families with recessive loss-of-function variants in the DRG1 translation factor. Detailed biochemical and functional analyses confirmed the pathogenicity of the variants in the novel developmental syndrome presented. Consistent with these variants driving the pathogenicity of the associated syndrome, we also document that *Drg1* is an essential gene in mice, where targeted inactivation causes preweaning lethality.

Our findings may predict the existence of a wider family of related neurodevelopmental disorders associated with pathogenic variants in genes encoding factors related to DRG1 biology. Considering the obligate nature of ZC3H15 for DRG1 function, and the common depletion of ZC3H15 to the DRG1 variants described here, one might predict the existence of a related neurodevelopmental disorder driven by pathogenic variants in this gene. Indeed, it has been reported that ZC3H15 has a similar pattern of tissue expression to DRG1, including in the developing central nervous system (Ishikawa *et al*, 2005), and the ZC3H15 gene is located within a chromosomal region altered in 2q32 deletion syndrome (OMIM612345, Van Buggenhout *et al*, 2005). Furthermore, the gene encoding the JMJD7 Jumonji-C oxygenase, which targets DRGs for lysyl hydroxylation (Markolovic *et al*, 2018), was identified as a candidate gene for autism and intellectual disability (Matsunami *et al*, 2014; de Ligt *et al*, 2012). Further work is required to fully understand the role of the JMJD7-DRG1/ZC3H15 pathway in cell biology and human disease (Westrip *et al*, 2021).

Although the precise molecular functions of the DRG1/ZC3H15 GTPase complex are still under debate, there is growing evidence supporting a fundamental role in translation (reviewed in Westrip *et al*, 2021), specifically the elongation step. Cryo-EM analyses of the yeast orthologues (Rbg1/Tma46) demonstrate associations with the A-site tRNA, the GTPase association centre, and the 40S subunit of the ribosome (Zeng *et al*, 2021). Precedence for the importance of translational elongation in neurodevelopment is underlined by other disorders driven by pathogenic variants in elongation factor pathways. For example, mutations in the eEF1 complex have also been implicated in developmental disorders associated with failure to thrive, developmental delay, intellectual disability, microcephaly, and facial dysmorphism (reviewed in McLachlan *et al*, 2019). Furthermore, mutations in the elongation factor EIF5A (MIM619376; Faundes-Banka Syndrome, Faundes *et al*, 2021) or an enzyme (Deoxyhypusine Synthase, MIM600944, Ganapathi *et al*, 2019) involved in its unique and essential modification, hypusination, have also recently been identified in neurodevelopmental disorders with clinical presentations that overlap with those described here. Interestingly however, aside from EEF1A2 (MIM602959, McLachlan *et al*, 2019), DRG1 represents the only other gene encoding a GTPase component of a translation elongation factor complex to have been identified thus far as the basis of a neurodevelopmental disorder. Our work also represents the first case of a disorder associated with the OBG/HflX GTPase family.

## MATERIALS AND METHODS

### Ethical Considerations

This study was approved in Oman by the ethics committees of the Medical Research Ethical Committee of the Sultan Qaboos University for Family 1, of KK Women’s and Children’s Hospital for Family 2, and Centogene (Germany) for Family 3. The parents of each family provided written informed consent to participate in this study and to publish their family pedigrees and clinical data. All clinical investigations were conducted according to the principles expressed in the Declaration of Helsinki. The study protocol was approved by A*STAR institutional review board (IRB 2019-087) and genetic analyses were performed in accordance with bioethics rules of national laws.

### Whole-Exome Sequencing

Exome sequencing was employed independently for the detection of variants in Families 1, 2 and 3. In brief, genomic DNA from peripheral blood sample was isolated using a DNeasy Blood and Tissue Kit (Qiagen, Courtaboeuf, France). DNA was barcoded and enriched for the coding exons of targeted genes using hybrid capture technology (Agilent–SureSelect Human All Exon). Prepared DNA libraries were then sequenced using Illumina paired-end Next Generation Sequencing (NGS) technology with average coverage of 100X. The reads were mapped against UCSC GRCh37/hg19 by Burrows-Wheeler Aligner and variants called using Genome Analysis Tool Kit. Variant filtration was conducted only to keep novel or rare variants (≤ 1%). Publicly available variant databases (1000 Genomes, Exome Variant Server, and gnomAD) and in-house exome databases were used to determine the frequency. Only coding/splicing variants were considered. The phenotype and mode of inheritance (autosomal recessive) were taken into account. The following criteria were then used to prioritise variants; high impact or highly damaging missense, a CADD score ≥ 20 and a variant within the autozygosity area. Sanger sequencing as standard was used to confirm the variants identified and segregation of the phenotype -genotype in the affected individuals.

### Isolation of human fibroblasts

Primary human cutaneous fibroblasts from the proband of Family 2 and one unaffected parental control were isolated from fresh skin biopsies. Briefly, biopsies were incubated in trypsin overnight at 4°C to enable peeling of the epidermis from the dermal compartment. Dermis was chopped up and stuck to a 10 cm plastic dish allowing the fibroblasts to migrate out of the dermal fragments.

### Cell culture

All HEK293T, HeLa, and fibroblast cells were cultured in DMEM supplemented with 10 % v/v Foetal Bovine Serum and 1 % v/v Pen/Strep, at 37°C, 5 % v/v CO_2_. Control primary fibroblasts used as WT controls were obtained from the SRIS Asian Skin Biobank with informed consent and prior IRB approval.

### Transfection

HEK293T and HeLa cells were transfected using FuGENE6 Transfection reagent (Promega). For a 15 cm plate 10 μg of DNA was added to 1 ml of OptiMEM and vortexed. 30 μl of FuGENE6 was then added and left to incubate at room temperature for 30 minutes before pipetting onto cells.

### Protein extraction

For pull down experiments, including for GTPase assays, cells were harvested 48 hrs after transfection, as follows. Cells were washed with cold PBS before lysis in 4 ml (for a 15 cm plate) of JIES buffer (100 mM NaCl, 20 mM Tris HCl pH 7.4, 5 mM MgCl_2_, 0.5 % (v/v) NP40) + 1x protease inhibitors (Sigma 58830). Protein lysates were centrifuged at 4°C for 10 minutes to pellet cell debris. For fibroblasts, cells were scraped in PBS and spun down. The cell pellets were then harvested in an appropriate volume of RIPA (150 mM NaCl, 5 mM EDTA, 50 mM Tris-HCl, 1% (v/v) NP-40, 0.5% Na Deoxychloride, 0.1% (w/v) SDS). Samples were normalised by measuring protein concentration using the Pierce 660 nm assay reagent (Thermo scientific), and diluting samples appropriately in lysis buffer. Western blot samples were boiled in Laemmli buffer for 5 minutes at 95° C.

### Immunoprecipitation

Protein extracts were immunoprecipitated with either anti-Flag M2 magnetic beads (Sigma, M8823) or anti-HA agarose beads (Sigma, A2095) at 4°C, overnight with rotation. To elute HA tagged DRG1 for western blotting, beads were washed 6 times in JIES buffer before boiling the beads in 2X Laemmli buffer for 5 minutes at 95°C.

### SDS PAGE western blot and ELISA

12 % (w/v) polyacrylamide gels were run at 150 volts in Tris Glycine SDS running buffer with Page Ruler Plus Protein Ladder (ThermoFisher). Proteins were transferred to a PVDF membrane (0.45 mm) at 320 milliamps for 25 minutes per membrane in Tris Glycine transfer buffer. The membrane was then blocked for 1 hour in 5 % (w/v) milk powder in PBS/0.1 % (v/v) Tween. The membrane was then incubated with one of the following primary antibodies: anti-ZC3H15 (Atlas Antibodies), anti-Flag HRP linked (Sigma), anti-β-actin HRP linked (Abcam), anti-V5 HRP linked (BioRad), or anti-HA HRP linked (Sigma). Anti-rabbit HRP conjugated secondary antibodies were used (Cell Signalling). Membranes were imaged using Clarity (Bio-Rad) or Femto (Thermo scientific) ECL blotting substrates using a Vilber Fusion Fx.

### Mouse mutant

The germline allele was generated at The Centre for Phenogenomics by electroporating Cas9 ribonucleoprotein complexes with single guide RNAs having spacer sequences of GAAAGGATCTTAGTCCAAGC targeting the 5’ side and TAAGAGTTACTATACTTGCC targeting the 3’ side of a critical region. This resulted in a 971-bp deletion Chr11:3262343-3263313_insA (GRCm38). Knockout mice were bred on a C57BL/6N background.

### GTPase assays

C-terminally Flag-tagged DRG1 was transiently expressed in HEK293Ts and immunoprecipitated as described above. Anti-flag immunoprecipitates were eluted for use in GTPase assays by incubating beads in GTPase buffer (100 mM Tris pH8, 300 mM KCl, 20 mM MgCl_2_, 10 % (v/v) glycerol) containing 150 mg/ml flag peptide, in a shaker at 1000 rpm for 30 minutes. GTPase activity of purified DRG1-Flag was measured using the Promega GTPase-Glo kit (V7681) as per the manufacturer’s instructions. The final concentration of GTP used was 5 mM. The final concentration of potassium ions used in the reaction was 150 mM, as previously described by others (Pérez-Arellano *et al*, 2013). The reactions were incubated overnight at 37°C. Luminescence was monitored with a PerkinElmer Enspire Multimode Plate reader. Each experiment was performed in triplicate.

### RNA extraction, reverse transcription and qPCR

Fibroblasts, seeded onto 10 cm plates, were scraped in PBS when 70-80% confluent. RNA was extracted from the cell pellets using a Sigma GeneElute Mammalian Total RNA extraction kit (RTN70). RNA quality was checked using an Agilent Qubit. RNA was then reverse transcribed using the Thermo High-Capacity cDNA Reverse Transcription kit (4368814) using the RNase inhibitor. The qPCR reactions were done using Fast Sybr Green Master Mix (Thermo, 4385612) as per the manufacturer’s instructions. Each biological repeat was performed in triplicate. GAPDH was used as the control. Primers are listed in Table S1. Reactions were run on a QuantStudio 5 Real-Time PCR System.

*DRG1* was amplified from cDNA prepared from fibroblasts using Phusion polymerase. The PCR reaction was run on a 1% (w/v) agarose gel and the band was cut out and the DNA extracted using a Sigma GeneElute Gel Extraction kit (NA1111). DNA samples were then sanger sequenced externally (Source Bioscience).

### Structural analyses

The crystal structure of Rbg1 (yeast DRG1) in complex with a C-terminal fragment of Tma46 (yeast ZC3H15) was used for the structural analysis, PDB code: 4A9A. Analysis was carried out using UCSF Chimera.

### Colony formation assays

Patient derived fibroblasts were seeded onto 10cm dishes at the stated density. Cells were left to grow for 10 days before staining with crystal violet. Images were taken using a Vilber Fusion Fx.

### Statistical analyses

Statistical analysis in Figure 2C, 2E, 2G, S2E-G and S3 was carried out using R.

## Data Availability

The data that support the findings of this study are available from the corresponding authors upon request.

## ACKNOWLEDGMENTS

We are grateful to all members of the Reversade, Al-Maawali and Coleman laboratories for support. M.L.C is a Cancer Research UK Fellow. B.R. is an investigator of the National Research Foundation (NRF, Singapore) and Branco Weiss Foundation (Switzerland) and an EMBO Young Investigator. We thank the SRIS Asian Skin Biobank (ASB), especially Alicia YAP Mei Yi, Joycelyn LEE Xiang Yi, and Siti Nur Aishah Binte ALIMAT, for isolating and expanding the primary fibroblasts. The ASB work was funded by the A*STAR IAF-PP Project (H1701a0004). This work was funded by a CRUK Programme Foundation Award to M.L.C. (C33483/A25674), a Strategic Positioning Fund for Genetic Orphan Diseases and an inaugural A*STAR Investigatorship from the Agency for Science, Technology and Research in Singapore to B.R. and the Singhealth Duke-NUS Genomic Medicine Centre Fund (SDDC/FY2021/EX/93-A147). F. P. is a recipient of a long-term European Molecular Biology Organization (EMBO) postdoc fellowship and a short-term EMBO travel fellowship. Her research is supported by the Singapore Ministry of Health’s National Medical Research Council under its Young Individual Research Grant scheme (Project ID MOH-000549-01) and A*STAR under its Career Development Award (Project number C210112002). A.A.M is a recipient of Sultan Qaboos University Strategic research funding (project code SR/MED/GENT/16/01).

## AUTHOR CONTRIBUTIONS

A.A.M., B.R., F.P., C.A.E.W., and M.L.C. designed the study. F.A.M, H.Q., B.C., E.S.T., M.N., S.A., I.L., G.F., A.A.M., and A.B.A. made clinical diagnoses and collected clinical data and samples. F.P., A.Y.J.N., C.B., B.V, B.R., A.B.A. and A.A.M. expanded patient cells and performed WES, homozygosity mapping, high throughput cohort re-sequencing and sequencing analyses. C.A.E.W. performed biochemical, structural and functional analyses of DRG1 patient variants. S.C.F., E.H., and U.B. contributed molecular biology. A.A.M., B.R., F.P., C.A.E.W., and M.L.C. wrote the manuscript with input from all co-authors.

## WEB RESOURCES

1000 Genomes Project Database, http://browser.1000genomes.org/index.html CRISPRScan, https://www.crisprscan.org

Exome Aggregation Consortium (ExAC), http://exac.broadinstitute.org

Exome Variant Server from NHLBI Exome Sequencing Project (ESP), https://evs.gs.washington.edu/EVS/

Genome Aggregation Database (GnomAD), http://gnomad.broadinstitute.org/ Greater Middle East (GME) Variome web, http://igm.ucsd.edu/gme/index.php NCBI dbSNP, https://www.ncbi.nlm.nih.gov/SNP/

Online Mendelian Inheritance in Man (OMIM), https://www.omim.org

