## Supplementary material for "Inactivation of DRG1, encoding a translation factor GTPase, causes a Recessive Neurodevelopmental Disorder": Figure S1

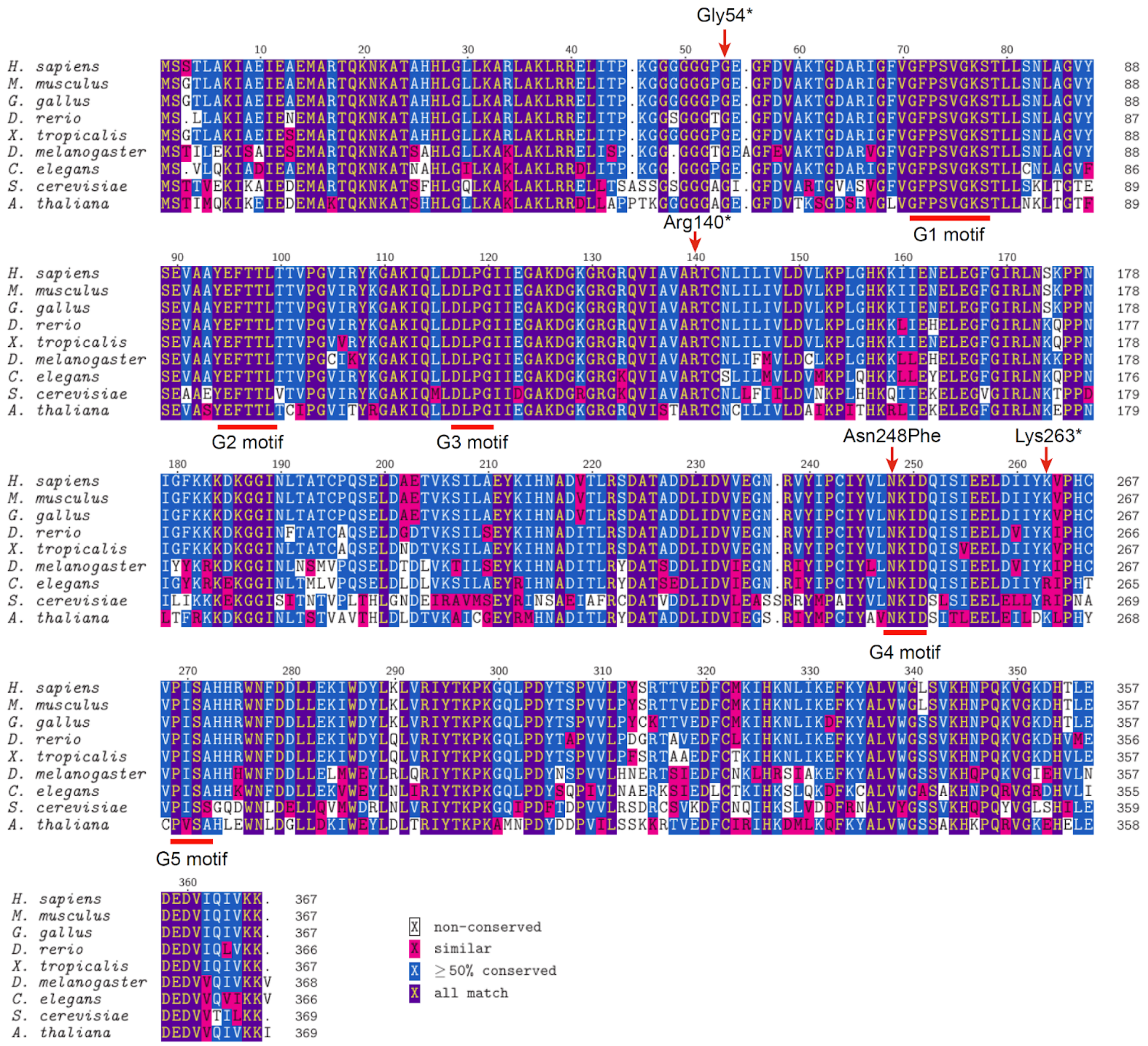


**Figure S1. Alignment of DRG1 homologues showing the location of the DRG1 disease associated variants.**

DRG1 protein sequences were aligned using ClustalW in MEGA7 (Kumar *et al*, 2016). The regions corresponding to the G motifs of the GTPase domain are indicated. See Table S2 for accession numbers for the sequences used.
