## Supplementary material for "Inactivation of DRG1, encoding a translation factor GTPase, causes a Recessive Neurodevelopmental Disorder": Figure S2

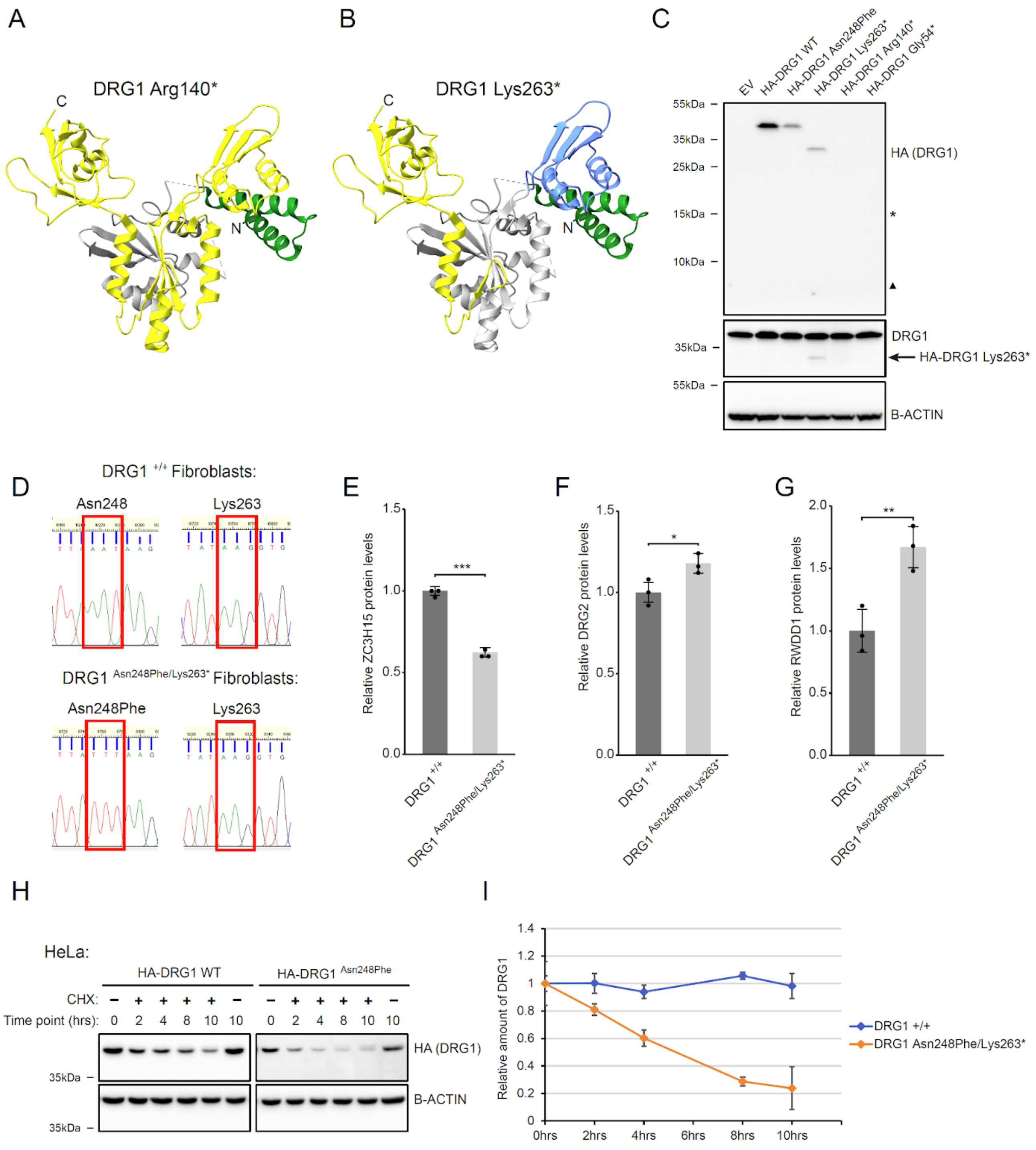


**Figure S2. Analysis of DRG1 variants expression and DRG1 Asn248Phe/Lys263* patient fibroblasts.**

A. Structural visualisation of the Arg140* truncation mutant. The region coloured yellow represents the part of DRG1 that would be absent from the protein. Image created in Chimera using the yeast Rbg1 structure from Francis *et al.*, (2012). Pdb: 4A9A.

B. Structural visualisation of DRG1 Lys263* same as in (A).

C. Western blots of lysates from HeLa cells expressing N-terminal HA-tagged DRG1 wildtype and variants. An empty vector (EV) pcDNA3 plasmid was used as a control. The asterisk and triangle indicate the predicted sizes of the Arg140* (15kDa) and Gly54* (7kDa) variants, respectively.

D. DRG1 sequencing traces showing the location of the Asn248Phe and Lys263* variants. DRG1 was PCR amplified from WT and DRG1 (Asn248Phe/Lys263*) cDNA (used in Figure 2C)) and sent for sequencing.

E-G. Quantification of ZC3H15, DRG2 and RWDD1 protein levels, respectively. Western blots using lysate from patient fibroblasts (Asn248Phe/Lys263*) were quantified using ImageJ. Data represents the mean of three repeats with error bars showing standard deviation. Statistical significance was estimated using a two-sample t-test. * = $p<0.05$, ** = $p<0.01$, *** = $p<0.001$.

H. Western blots from a cycloheximide (CHX) stability assay of HA-DRG1 WT or Asn248Phe transiently expressed in HeLa cells. Cells were treated with 50µg/ml CHX and then harvested at the indicated time points. A 10hrs dimethyl sulfoxide (DMSO) control was also included.

I. Quantification of DRG1 protein levels from fibroblast (Asn248Phe/Lys263*) CHX stability assay (Figure 2F). Data represents the mean of 2 repeats with error bars showing standard deviation.
