## Supplementary material for "Inactivation of DRG1, encoding a translation factor GTPase, causes a Recessive Neurodevelopmental Disorder": Figure S3

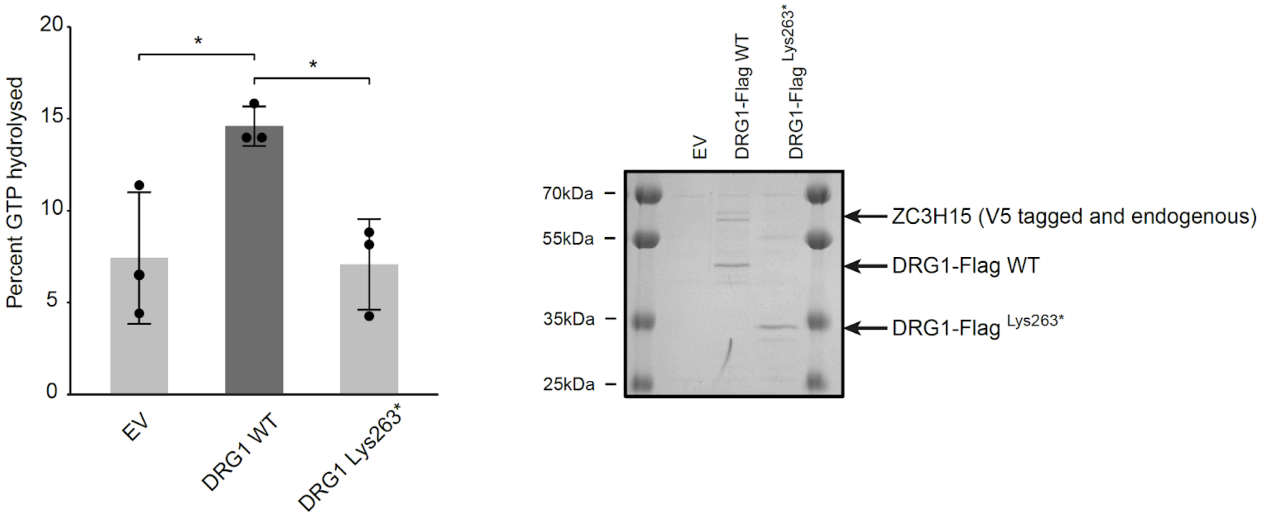


**Figure S3. DRG1 Lys263* has reduced GTPase activity.**

The left panel shows the results of a GTPase assay using C-terminally flag tagged DRG1 WT and Lys263* that were co-transfected with C-terminally V5 tagged ZC3H15 in HEK293T cells and purified using anti-flag pulldown. The data represents the mean with error bars showing the standard deviation of n=3 biological repeats (data points shown). Statistical significance was confirmed using a one-way ANOVA with Tukey HSD to estimate p values. Right panel shows a representative Coomassie-stained gel of the purified DRG1/ZC3H15 proteins.
