## Supplementary material for "Inactivation of DRG1, encoding a translation factor GTPase, causes a Recessive Neurodevelopmental Disorder": Table S1

**Table S1. Primers used for qPCR.**

| Gene of interest | Sequence (5'-3') |
|---|---|
| *DRG1* | Forward: GATGTGGTGGAAGGAAACAGA<br>Reverse: GTACACAGTGAGGCACCTTATAG |
| *GAPDH* | Forward: AGCCACATCGCTCAGACAC<br>Reverse: GCCCAATACGACCAAATCC |
