## Supplementary material for "Inactivation of DRG1, encoding a translation factor GTPase, causes a Recessive Neurodevelopmental Disorder": Table S2

**Table S2. Accession numbers for sequences used in DRG1 alignments.**

| **Species** | **Protein name** | **Accession number** |
|---|---|---|
| *Homo sapiens* | DRG1 | AAH20803.1 |
| *Mus musculus* | DRG1 | NP_031905.1 |
| *Gallus* | DRG1 | NP_001027533.1 |
| *Danio rerio* | DRG1 | NP_956332.1 |
| *Xenopus tropicalis* | DRG1 | AAH80366.1 |
| *Drosophila melanogaster* | DRG1 | NP_536733.1 |
| *Caenorhabditis elegans* | DRG1 | NP_001255126.1 |
| *Saccharomyces cerevisiae* | DRG1 | AJP36937.1 |
| *Arabidopsis thaliana* | DRG1 | NP_195662.1 |
